# The valuation of outcomes for the temporary and chronic health states associated with Chlamydia trachomatis infection

**DOI:** 10.1101/2021.06.15.21258547

**Authors:** Chidubem B.Okeke Ogwulu, Louise J. Jackson, Philip Kinghorn, Tracy E. Roberts

**Author notes:** Corresponding author: Tracy Roberts, Professor of Health Economics, Health Economics Unit, Institute of Applied Health Research, College of Medical and Dental Sciences, IOEM Building, University of Birmingham, Edgbaston, Birmingham, B15 2TT, UK. Funding source: This study was conducted as part of a University of Birmingham, College of Medicine and Dental Sciences Studentship.

## Abstract

**Background:** Eliciting health-state utility values (HSUVs) for some diseases is complicated by the mix of associated temporary (THSs) and chronic health states (CHSs). This study uses one such disease, chlamydia infection, to explore the challenges. The objectives were to:

1. Define a set of health-state descriptions related to chlamydia and
2. Derive HSUVs for these health states (both temporary and chronic).

**Methods:** HSUVs were elicited for seven health states (five THSs and two CHSs) depicting the symptoms of chlamydia, developed using evidence from the literature and clinical experts. The chained time trade-off (TTO) and visual analogue scale (VAS) were applied to THSs while conventional TTO was applied to the CHSs. Ectopic pregnancy was used as the anchor for the chained TTO approach. The study sampled from three different population groups and the survey was administered face-to-face.

**Results:** One hundred participants were assessed with an interview completion rate of 100%. Mean TTO utilities were consistently higher than VAS scores. The aggregated mean chained TTO results for the THS ranged from 0.46 (SD, 0.24) for ectopic pregnancy, to 0.77 (SD 0.21) for cervicitis.

**Conclusions:** Chained TTO was shown to be feasible in this population and the resulting HSUVs could have implications for economic evaluations for chlamydia prevention and control. Methodological challenges included the development of health-state descriptions, the selection, and the duration of appropriate anchor state.

## INTRODUCTION

Economic evaluations informing decisions made by the UK’s National Institute for Health and Care Excellence (NICE) are recommended to use cost-utility analysis (CUAs), with results reported in terms of cost per quality-adjusted life-years (QALYs) [1]. The QALY is a composite index of the quantity and quality of life (QoL). While the quantity of life is an accepted conventional measure [2], QoL estimation is sometimes both challenging and controversial [2,3]. To weight changes in life expectancy due to QoL experienced, QALYs use health utilities which are measures of the strength of preferences that people have for specific health-states [4]. Two groups of methods exist for measuring utilities, direct and indirect methods [2,3].

For direct methods, respondents typically value hypothetical health-states and preferences are directly measured, using time trade-off (TTO), standard gamble (SG) or visual analogue scale (VAS) technique [3]. For indirect methods, respondents complete generic health-related QoL (HR-QoL) questionnaires such as the EQ-5D and preferences are indirectly mapped using pre-valued algorithms [5]. The indirect methods instruments are available ‘off-the-shelf’, user friendly and circumvent the time-consuming exercise of asking respondents to trade health-states for various risks of death (or years of life remaining) each time [6].

Despite the strengths of indirect methods, NICE recognises that for some situations, the methods are unsuitable, or data are unavailable for the patient population [7] and recommends a direct choice-based technique such as TTO or SG [7] for such situations. In this study, the disease focus (Chlamydia trachomatis infection) makes the direct approach more suitable, primarily because the disease is typically asymptomatic and may have multiple impacts occurring at different times [8]. Henceforth this paper will focus on direct valuation approaches.

The calculation of QALYs involves the determination and definition of relevant health-states [9] followed by the valuation of each health-state to assign health-state utility values (HSUVs) [9,10]. Selecting an appropriate valuation technique is often challenging [3], more so if the condition is associated with temporary health-states (THSs) which may require special consideration of methodological issues [10].

Unlike the valuation techniques for chronic health-states (CHSs) which are widely documented [11,12] the techniques for THSs have received less attention, with no guiding principle. Conventional techniques such as TTO and SG are deemed inappropriate for THSs as they are inherently tailored for chronic durations [13]. Techniques that are *adapted* from conventional methods such as chained approaches have been proposed specifically for THS valuation and incorporate the acuity of certain health-states [14].

A previous paper [8] outlined the challenges in HSUV elicitation for some diseases which included the variation in the duration of the health-states since some sexually transmitted infections (STIs) involve both temporary and CHSs. The current paper explores this challenge.

## BACKGROUND

In this paper, our methodological exploration uses Chlamydia trachomatis infection (hereon referred to as chlamydia) as a case study. Chlamydia is common [15] and most prevalent in sexually active young adults aged 15 to 24 years (3-6%) [16]. Chlamydia is mostly asymptomatic at onset and if undetected, can progress to complications like pelvic inflammatory disease (PID) in women [17,18]. Yet chlamydia is curable if detected, hence, in many developed countries screening is recommended for asymptomatic sexually active young adults [19-21]. Chlamydia imposes a considerable financial burden on health services in many countries with the English National Chlamydia Screening Programme (NCSP) [19] costing over £100 million annually [22].

Few of the previous economic evaluations of chlamydia interventions presented outcomes in terms of QALYs [8,23]. Evaluations that have reported QALYs provided little information on how these QALYs were derived (where indirect methods were used) or on the conceptualisation of the health-states, and the derivation of HSUVs (where direct methods were used) [8,23]. However, if the QALY estimations used in these evaluations are not robust then the associated policy recommendations may not be optimal.

This study aimed to estimate the HSUVs associated with chlamydia using direct methods. The choice of a direct methods approach was because instruments such as the EQ-5D which are the mainstay of the indirect approach were not considered appropriate for THS valuation [14,24]. The argument for using the direct approach is mostly related to the difficulty in accessing robust primary data for the disease and its complications [8]. To use an indirect approach, one would need to capture data from different patients depicting the range of different health-states at different time points [11]. Additionally, some complications of chlamydia such as infertility and PID which occur in the future are also associated with other causes such as gonorrhoea and cancer treatments [8]. Hence, if patients develop these complications their relation to chlamydia or other causes is unknown. Prospective data collection from individuals diagnosed with chlamydia would require the denial of treatment to such patients, which is unethical [15,16].

The objectives of the study were:

1. To define health-state descriptions related to chlamydia in women
2. To elicit utilities for the THSs and CHSs associated with chlamydia.

A systematic review of the methodological approaches for valuing THSs is reported elsewhere [25]. Based on the review’s evidence, conventional TTO was selected for the CHSs and chained TTO for the THSs.

Chained TTO is a two-stage procedure for measuring THS utilities [26]. Although considered appropriate by many authors for valuing THSs, chained TTO is often avoided as it is perceived as difficult and cognitively burdensome for respondents [25,27,28]. An additional aim of the study was to explore the feasibility of the chained TTO approach.

## METHODS

### Health-state Descriptions

Health-state descriptions were developed in three steps: literature review [15,17], use of expert opinion and a pragmatic review by yound adults for clarity (Supplemental_1). Seven health-state scenarios were developed and included (i) Cervicitis (ii) Mild pelvic inflammatory disease (PID), (iii) Severe PID, (iv) Ectopic pregnancy, (v) Pre-term delivery, (vi) Chronic pelvic pain, and (vii) Tubal factor infertility (TFI). The first five health-states are considered THSs and the last two, CHSs. Each scenario described a health-state in terms of the symptoms, impact on health-related QoL, treatment, and implications (Supplemental_2).

### Recruitment and Data Collection

To be included in the study, participants had to be aged ≥16 year, and be able to read and understand English. To reflect NICE guidance on the source of preference data [1] (which requires the use of general public values), data were collected separately from three groups using the same valuation technique.

The first group comprised undergraduates at a UK University. This was primarily a convenience sample and the aim was to test the feasibility and identify any methodological issues. The second group was individuals attending a Sexual Health Clinic (SHC) in a metropolitan area of the West Midlands, which also offered family planning services. These individuals could be patients with an STI, at risk of having an STI or seeking advice about sexual health or contraception more generally. Individuals will have been attending the clinic for a variety of reasons, so it is not appropriate to think about this group as a homogenous group of patients attending for treatment. Ethical approval was obtained from the NHS Health Research Authority (IRAS ID-194331). The third group was a convenience sample of staff and PhD students from the authors’ academic institute. Ethical approval for Group 1 and 3 was obtained from the appropriate University body (ERN_16-1522).

All surveys were interviewer-administered. Following the study’s introduction by the researcher, individuals who expressed willingness to participate were asked to sign a consent form before the questionnaire administration. Respondents at the SHC were given £5 gift vouchers as a thank you.

### Valuation

The study questionnaire (Supplemental_3) is divided into five sections (A-E) - the first four sections comprised health-state valuation exercises, while the last section contained questions covering background information. Each section included an example as guidance on how to complete that section. To avoid respondents’ bias, the examples for each section did not relate to sexual health. The background questions were selected from the British National Surveys of Sexual Attitudes and Lifestyles (NATSAL) [29].

### Visual Analogue Scale

The VAS was applied to the THSs. Using a vertical line 10 cm in length, anchored at zero (0) by worst imaginable health and at 100 by best imaginable health, participants were asked to indicate how good or bad a health-state would be for them. Dead was not used as the lower anchor because of the temporary nature of the health-states. To reduce respondents’ burden, each participant was randomised to rank only three out of the five THSs.

### Conventional TTO

Conventional TTO [26] was applied to the CHSs - chronic pelvic pain and TFI. Using a TTO table, participants were asked to choose between living in a CHS for 30 years (**t**) and then dying and living in full health for a period (**x**), shorter than **t**. The time (**x**) in full health was varied in intervals of one to five years until respondents expressed their indifference between the alternatives. A 30-year duration was selected based on the nature of the health-states, participants’ life expectancies and previous studies [30,31]. The utility for the CHS was calculated as **x/t** [26].

The TTO table format (Figure 1) was based on the self-completion method of conventional TTO, adapted from the York Measurement and Valuation of Health (MVH) TTO guide for TTO without props [32]. However, to ensure an easier procedure for both the researcher and the participants, minor modifications were made to the table format as used by previous studies [33, 34].

**Figure 1.**
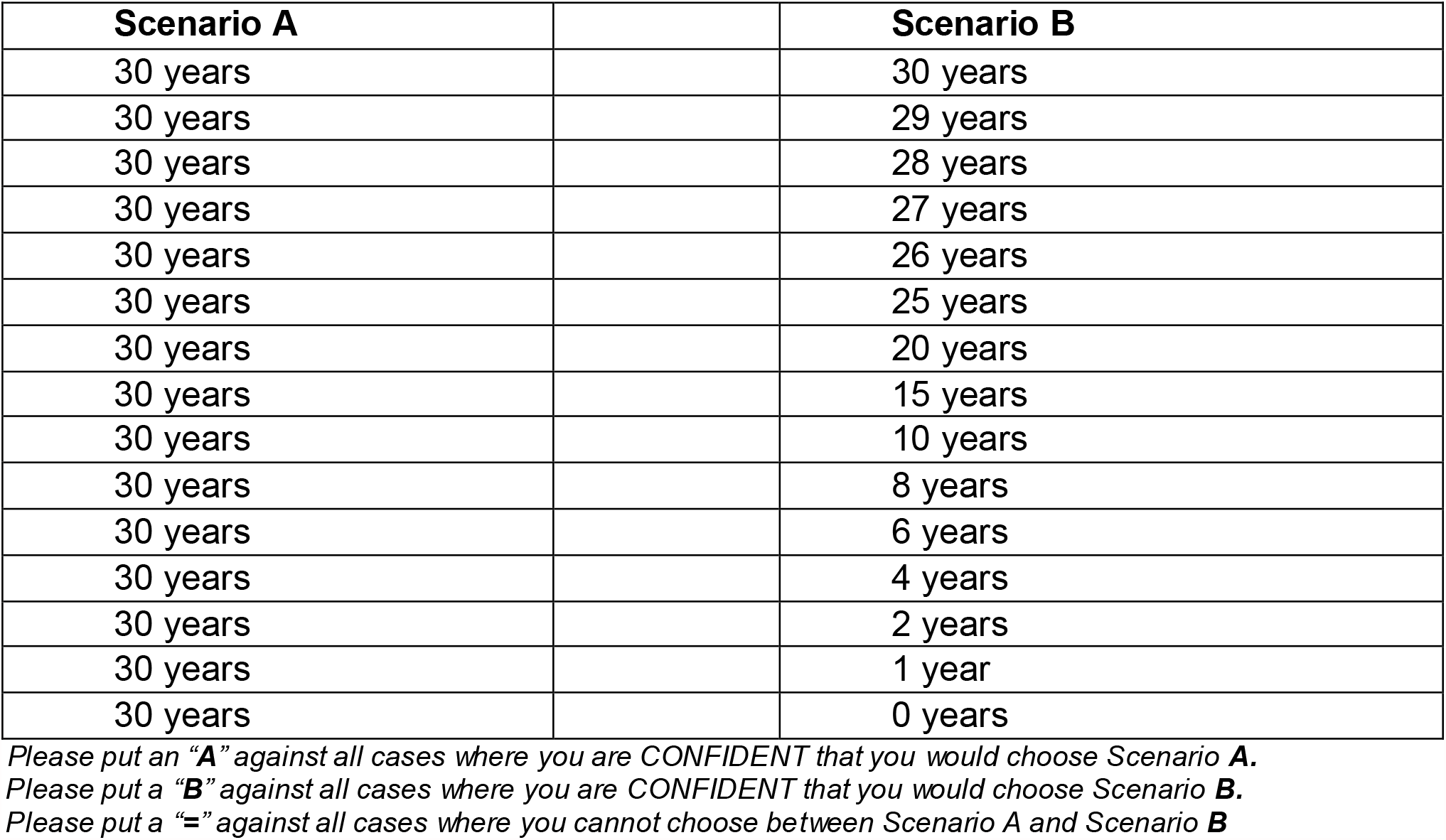
The Time Trade-off Table.

### Chained TTO

Chained TTO was applied to five THSs. The chained TTO procedure is a two-stage process for measuring THS utilities which can be applied to conventional TTO, SG or VAS procedures [11]. In addition to the standard anchor points of Full health and Dead (used in conventional methods), the chained procedure uses additional health-states (depicting the anchor health-state) as endpoints. The anchor health-state is typically the worst THS from the set of THSs being valued and is ideally worse than the THS being assessed but better than Dead [11].

In Stage-I of chained TTO (Figure 2a), a THS (**h**) was valued relative to full health (**1**) and the anchor health-state (**w**). Participants were asked to choose between two scenarios. Either the THS (**h**) for 12 months (**t**_**h**_) (Scenario A), or the anchor health-state (**w**) for a duration **x**_**w**_, less than **t**_**h**_, followed by a return to full health (Scenario B). The time **x**_**w**_ was varied until the participant expressed indifference for either scenario. The THS score at this stage was calculated as **x**_**w**_**/t**_**h**_ indicating the value of the THS (**h**) relative to the anchor health-state and full health.

**Figure 2a.**
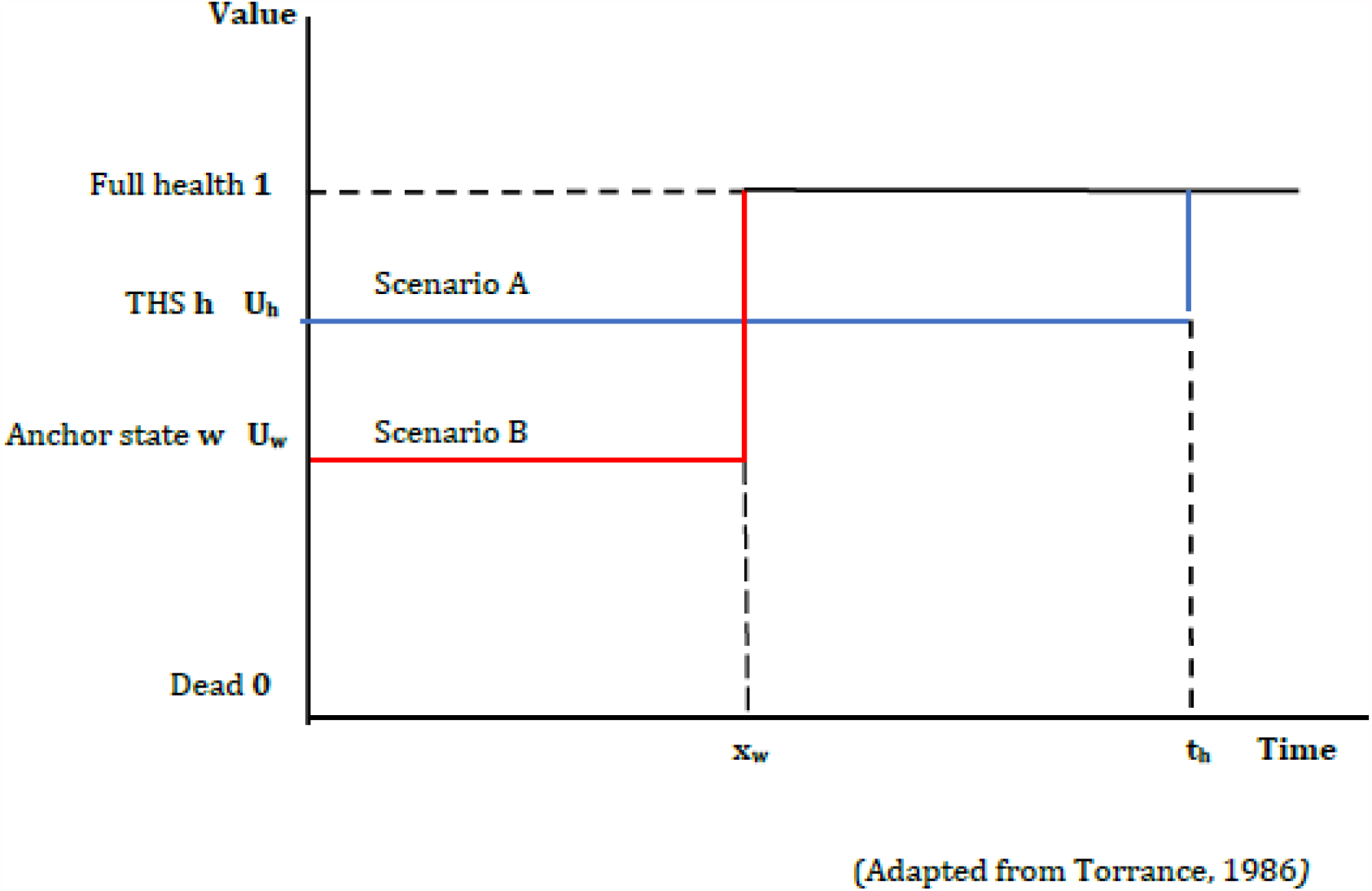
Stage-I of the Chained Time Trade-off.

In Stage-II (Figure 2b), the anchor health-state (**w**) (Scenario A) was compared with Full health (**1**) and Dead (**0**) for a 10-year duration (**x**_**1**_) to derive the HSUV (**U**_**w**_) for the anchor health-state using **x**_**1**_**/t**_**w**_ [11]. With this approach, the THS is valued indirectly against full health and dead using an ‘intermediate’ anchor health-state, followed by a return to full health.

**Figure 2b.**
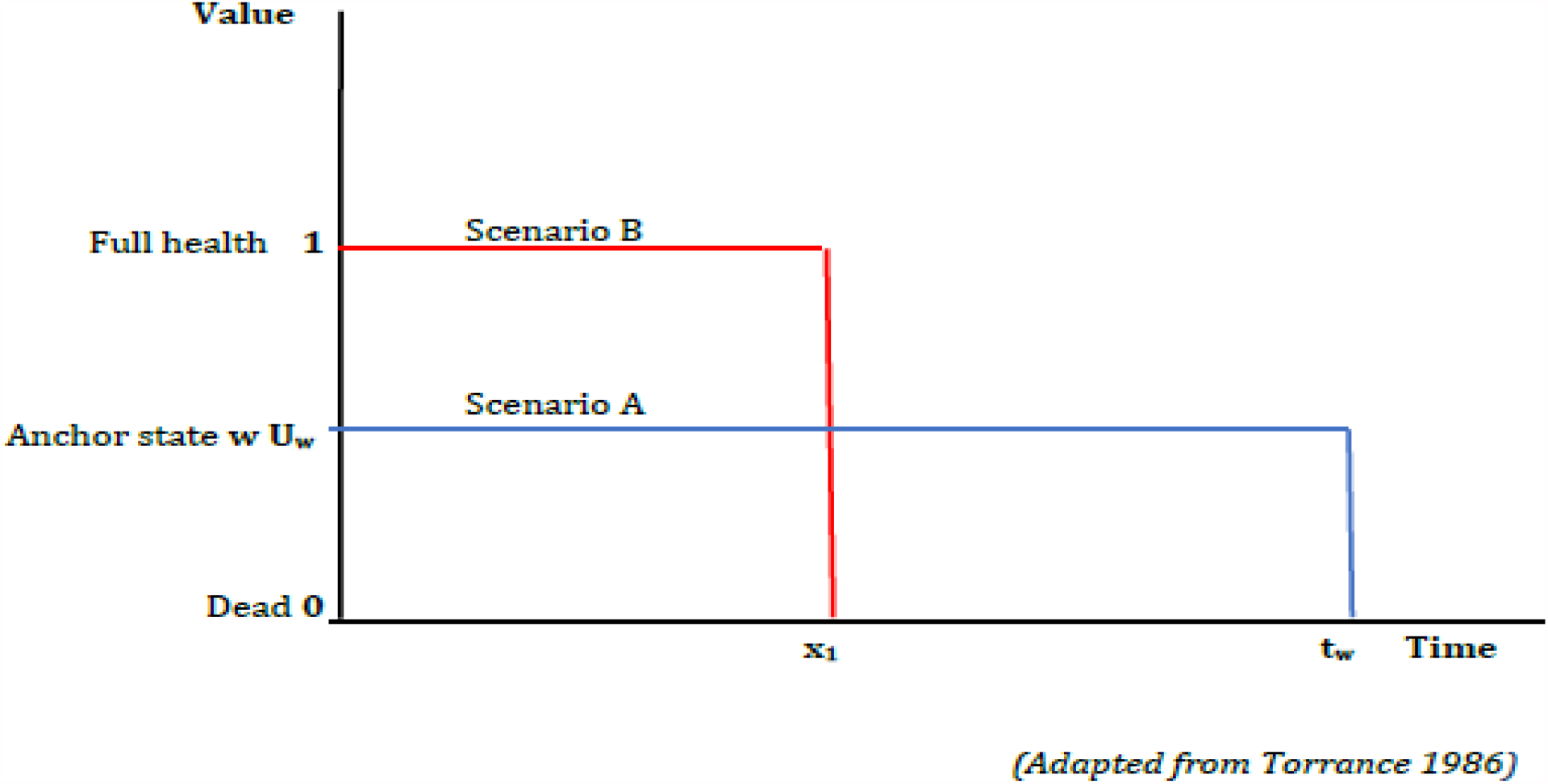
Stage-II of the Chained Time Trade-off.

The utility of the THS from Stage-I (**U**_**h**_) was calculated by transforming the value for Stage-I from the ‘anchor health (<1) to full health (1)’ scale, to the ‘dead (0) to full health (1)’ scale using the formula

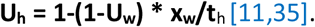

In cases, where participants preferred the anchor health-state to another THS, as per the recommendation by Jansen and colleagues [36], rather than shortening the time in the anchor state, the time was extended in increments of 3 months until a point of indifference was reached. For example, participants were asked to choose between 15 months in the THS and 12 months in the anchor state, then 18 months in the THS or 12 months in the anchor and so on.

### Duration

The paucity of studies on chained TTO means that there are no guidelines for operationalisation. In proposing chained TTO, Torrance [11] specified that a temporary duration is used for Stage-I. A temporary duration is defined as lasting for 12 months or less [25]; thus we used a 12-month duration for this stage. However, for Stage-II, Torrance stated that the anchor health-state should be valued as a short-duration chronic state; this has been echoed by other authors [10,35]. However, there is no clear definition of what period constitutes a short-term chronic duration. A targeted review of papers that have used chained TTO showed that studies used different durations to value the anchor health-state. After consideration of the review findings, the disease condition, and respondents’ ages, we selected 10 years as the anchor health-state duration. The rationale for this would be that for the chained approach, the duration of the anchor HS is typically less than the duration of the chronic HS

### Statistical Analysis

Data analysis was carried out using STATA 13.0 [37]. Summary statistics were generated for the descriptive variables, VAS scores, and TTO utilities. Consistency between chained TTO and VAS was assessed by analysing repondents’ rank order for the THSs at face value. Differences in mean values between the groups were compared using the one-way analysis of variance (ANOVA) test [38] and post-hoc tests (the Tukey’s HSD (honestly significant difference)). Correlation between the different techniques was examined using Pearson’s correlation test [38]. Regression analysis was used to examine associations between VAS/TTO results and respondents’ age group and gender. Additionally, the feasibility of the techniques was reported based on criteria used by previous studies such as the number of completed interviews and missing answers [35,39,40].

## RESULTS

One hundred individuals including 37 students, 37 SHC attendees, and 26 University Staff members took part in this study. Following the valuation of the students (Group-1), minor modifications were made to the health-state descriptions texts before interviewing the other groups. A 100% questionnaire/interview completion rate was achieved with an average completion time range of 21-31 minutes. There were no missing values.

Respondents’ characteristics are presented in Table 1. Respondents included 58 females, 41 males and one person who identified as non-binary. The modal age groups were 20-24 years for Group-1, 16-17 years/20-24 years for Group-2, and 25-34 years for Group-3. Just over half (n = 51) of the participants were of White ethnicity and 30 participants were Black.

**Table 1:** Descriptive characteristics for Study Respondents.

| **Variable** | **n (%)**<br>**Students**<br>**(Group-1)**<br>**N = 37** | **n (%) SHC**<br>**attendees (Group-**<br>**2)**<br>**N = 37** | **n (%) University**<br>**Staff**<br>**(Group-3)**<br>**N = 26** | **Total**<br>**N = 100** |
| --- | --- | --- | --- | --- |
| **GENDER** |  |  |  |  |
| Female | 24 (64.86) | 20 (54.05) | 14 (53.85) | 58 (58.00) |
| Male | 12 (32.43) | 17 (45.95) | 12 (46.15) | 41 (42.00) |
| Other | 1 (2.70) |  |  | 1 (1.00) |
| **ETHNICITY** |  |  |  |  |
| Asian/Asian British | 8 (21.62) | - | 3 (11.54) | 11 (11.00) |
| Black British | 1 (2.70) | 8 (21.62) | 3 (11.54) | 12 (12.00) |
| Black, other background | 1 (2.70) | 12 (32.43) | 5 (19.23) | 18 (18.00) |
| Mixed ethnic group | 3 (8.11) | 1 (2.70) | - | 4 (4.00) |
| Other, ethnic group | 1 (2.70) | - | 1 (3.85) | 2 (2.00) |
| White British | 21 (56.76) | 16 (43.24) | 7 (26.92) | 44 (44.00) |
| White, other background | 2 (5.41) | - | 7 (26.92) | 7 (7.00) |
| **AGE AT LAST BIRTHDAY** |  |  |  |  |
| 16-17 | - | 11 (30.56) | - | 11 (11.00) |
| 18-19 | 1(2.70) | 5 (13.89) | - | 6 (6.00) |
| 20-24 | 36 (97.30) | 11 (30.56) | - | 47 (47.00) |
| 25-34 | - | 8 (22.22) | 15 (57.69) | 23 (23.00) |
| 35 or older | - | 1 (2.78) | 11 (42.31) | 12 (12.00) |
| **EMPLOYMENT** |  |  |  |  |
| Employed/self-employed | - | 9 (24.32) | 15 (57.69) | 24 (24.00) |
| Seeking work | - | 6 (16.22) | 1 (3.85) | 7 (7.00) |
| Student | 37 (100) | 20 (54.55) | 10 (38.46) | 67 (67.00) |
| Others | - | 2 (5.41) | - | 2 (2.00) |

### Visual Analogue Scale

For respondents’ VAS scores, out of a possible score of 0-1, the lowest mean value was assigned to ectopic pregnancy (0.30, SD-0.19) and the highest was to cervicitis (0.51, SD-0.22) (Table 2). Ectopic pregnancy score was slightly lower than severe PID score (0.33, SD-0.23). The interquartile ranges (IQR) for the median values were small, indicating considerable consistency among respondents’ valuations.

**Table 2:** Mean and Median VAS* and TTO values

| Health-state | Group-1<br>n=37 | Group-2<br>n=37 | Group-3<br>n=26 | Mean total<br>n=100 |
| --- | --- | --- | --- | --- |
| CERVICITIS |  |  |  |  |
| Mean TTO (SD) | 0.69 (0.25) | 0.77 (0.18) | 0.90 (0.11) | 0.77 (0.21) |
| Median TTO (IQR) | 0.67 (0.50-0.91) | 0.79 (0.71-0.86) | 0.92 (0.83-1) | 0.81 (0.65- 0.95) |
| Mean VAS (SD) | 0.47 (0.20) | 0.45 (0.23) | 0.64 (0.18) | 0.51 (0.22) |
| Median VAS (IQR) | 0.45 (0.31-0.58) | 0.40 (0.30-0.60) | 0.61 (0.55-0.70) | 0.5 (0.31-0.70) |
| MILD PID |  |  |  |  |
| Mean TTO (SD) | 0.74 (0.23) | 0.76 (0.20) | 0.82 (0.15) | 0.76 (0.20) |
| Median TTO (IQR) | 0.76 (0.55-1.00) | 0.81 (0.70-0.87) | 0.86 (0.75-0.92) | 0.82 (0.61-0.94) |
| Mean VAS (SD) | 0.40 (0.19) | 0.34 (0.25) | 0.49 (0.18) | 0.40 (0.22) |
| Median VAS (IQR) | 0.35 (0.25- 0.57) | 0.27 (0.20- 0.40) | 0.50 (0.40-0.60) | 0.37 (0.24-0.52) |
| SEVERE PID |  |  |  |  |
| Mean TTO (SD) | 0.81 (0.27) | 0.75 (0.17) | 0.61 (0.27) | 0.74 (0.24) |
| Median TTO (IQR) | 0.97 (0.60-1.00) | 0.77 (0.65-0.86) | 0.68 (0.45-0.79) | 0.65 (0.60-0.96) |
| Mean VAS (SD) | 0.31 (0.24) | 0.32 (0.27) | 0.38 (0.17) | 0.33 (0.23) |
| Median VAS (IQR) | 0.25 (0.15-0.40) | 0.28 (0.10-0.50) | 0.32 (0.25-0.41) | 0.30 (0.15-0.41) |
| ECTOPIC PREGNANCY |  |  |  |  |
| Mean TTO (SD) | 0.47 (0.24) | 0.35 (0.24) | 0.58 (0.17) | 0.46 (0.24) |
| Median TTO (IQR) | 0.50 (0.30-0.60) | 0.35 (0.20-0.50) | 0.58 (0.45-0.70) | 0.45 (0.30-0.60) |
| Mean VAS (SD) | 0.30 (0.18) | 0.26 (0.23) | 0.36 (0.16) | 0.30 (0.19) |
| Median VAS (IQR) | 0.25 (0.19-0.39) | 0.21 (0.10-0.38) | 0.35 (0.26-0.45) | 0.27 (0.15-0.40) |
| PRE-TERM DELIVERY |  |  |  |  |
| Mean TTO (SD) | 0.77 (0.24) | 0.74 (0.26) | 0.76 (0.30) | 0.76 (0.26) |
| Median TTO (IQR) | 0.87 (0.60-1.00) | 0.78 (0.68-0.91) | 0.88 (0.75-0.95) | 0.85 (0.6-0.99) |
| Mean VAS (SD) | 0.32 (0.18) | 0.46 (0.22) | 0.48 (0.22) | 0.41 (0.21) |
| Median VAS (IQR) | 0.30 (0.20-0.47) | 0.44 (0.33-0.65) | 0.50 (0.30-0.61) | 0.40 (0.30-0.60) |
| CHRONIC PELVIC PAIN |  |  |  |  |
| Mean TTO (SD) | 0.38 (0.27) | 0.31 (0.27) | 0.56 (0.20) | 0.40 (0.27) |
| Median TTO (IQR) | 0.30 (0.20-0.50) | 0.27 (0.10-0.40) | 0.50 (0.5-0.67) | 0.33 (0.2-0.67) |
| Mean VAS (SD) | N/a | N/a | N/a |  |
| Median VAS (IQR) | N/a | N/a | N/a |  |
| TUBAL FACTOR INFERTILITY |  |  |  |  |
| Mean TTO (SD) | 0.50 (0.33) | 0.44 (0.31) | 0.75 (0.20) | 0.54 (0.32) |
| Median TTO (IQR) | 0.67 (0.20-0.67) | 0.4 (0.20-0.67) | 0.83 (0.67-0.9) | 0.67 (0.30-0.83) |
| Mean VAS (SD) | N/a | N/a | N/a |  |
| Median VAS (IQR) | N/a | N/a | N/a |  |

\*The VAS values were converted from a 0-100 to a 0-1 scale by dividing each score by 100.

### Time Trade-off

The mean utilities for the THSs obtained for chained TTO (Table 2 and Figure 3) were, in ascending order: ectopic pregnancy 0.46 (SD-0.24), severe PID 0.74 (SD-0.24), pre-term delivery 0.76 (SD-0.26), mild PID 0.76 (SD-0.20) and cervicitis 0.77 (SD-0.21).

**Figure 3.**
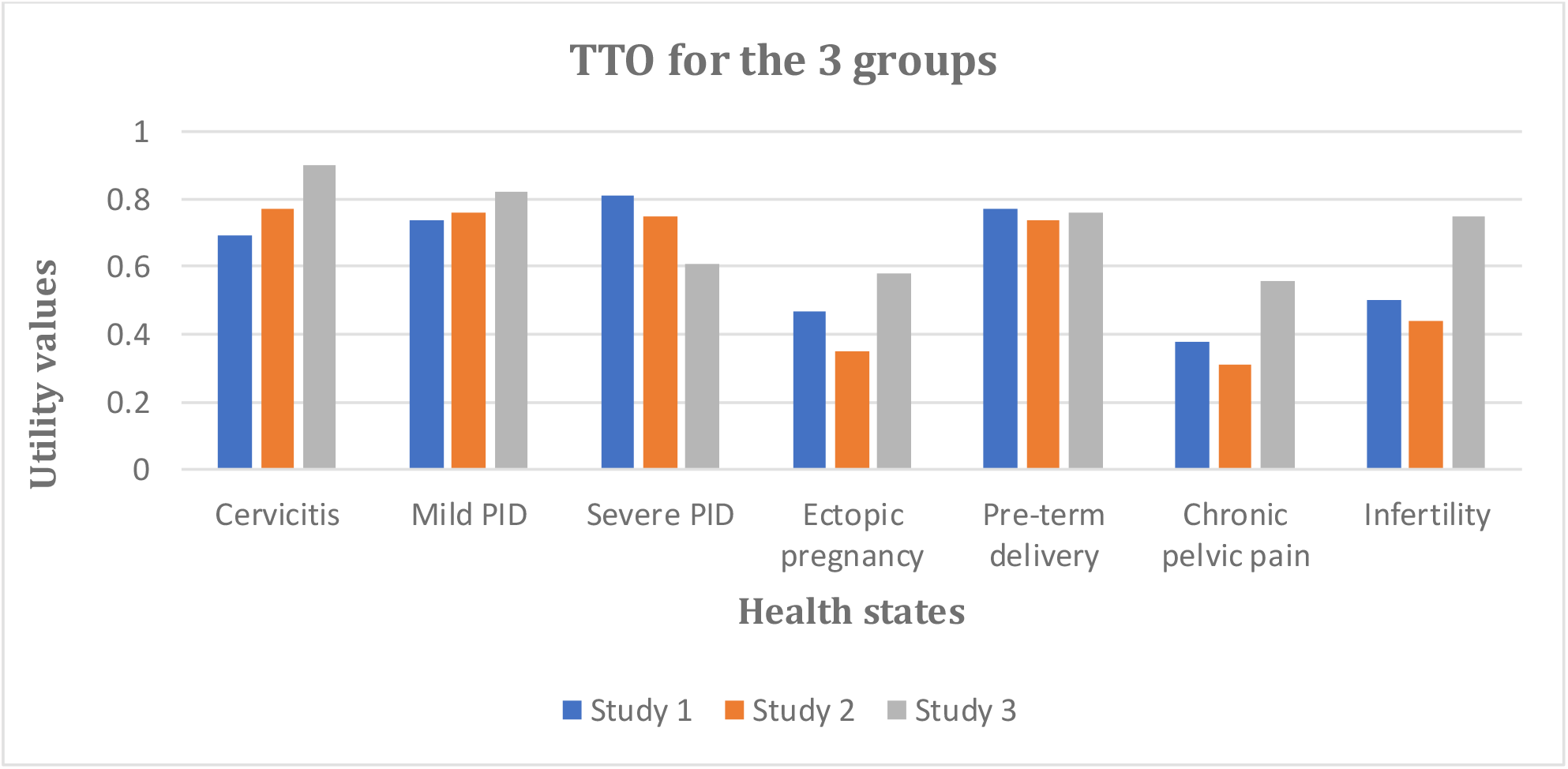
TTO values for the three groups.

The rank order of health-states for VAS and chained TTO techniques followed the expected order from clinical evidence [18, 41]. The mean value of 0.46 assigned to ectopic pregnancy was relatively low compared to those for other THSs, which ranged from 0.74 to 0.77. For the CHSs, the mean HSUVs were 0.40 (SD-0.27) for chronic pelvic pain and 0.54 (SD-0.32) for TFI.

### Comparison by Group

Table 3 presents a summary of the ANOVA and post-hoc Tukey results. The difference in mean VAS scores was statistically significant for cervicitis (*F*_*(2, 62)*_*:4*.*97, p:0*.*0099*) only. The post-hoc test for this health-state revealed that the scores were statistically significantly higher in Group-3 than in Group 1 (0.168 ± 0.065, p:0.032) and 2 (0.192 ± 0.064, p:0.011). Post-hoc tests showed varying statistical significance between groups (Table 3).

**Table 3:**
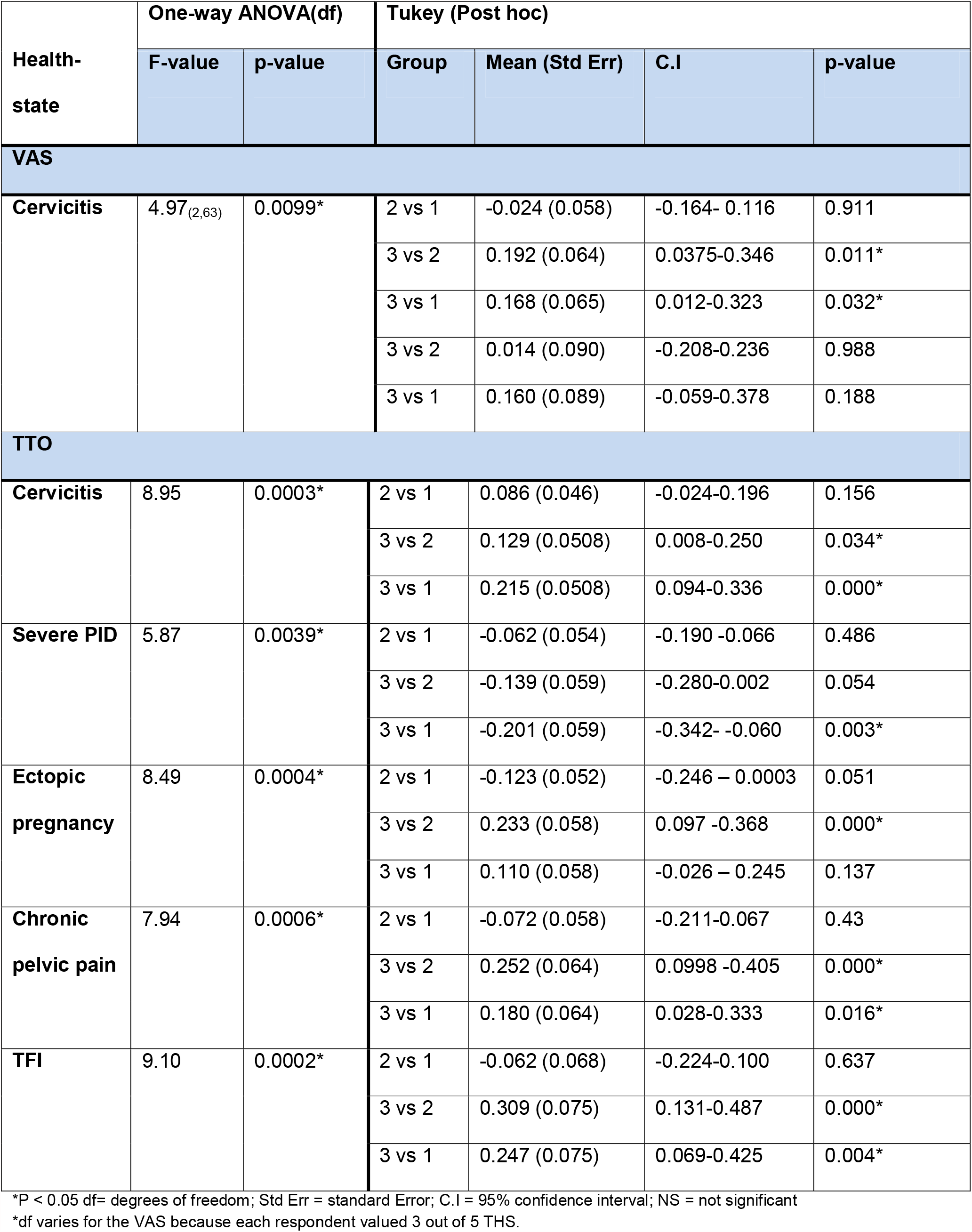
One-way ANOVA for mean differences

|  | One-way ANOVA(df) |  | Tukey (Post hoc) |  |  |  |
| --- | --- | --- | --- | --- | --- | --- |
| Health-state | F-value | p-value | Group | Mean (Std Err) | C.I | p-value |
| VAS |  |  |  |  |  |  |
| Cervicitis | 4.97<sub>(2,63)</sub> | 0.0099* | 2 vs 1 | -0.024 (0.058) | -0.164- 0.116 | 0.911 |
|  |  |  | 3 vs 2 | 0.192 (0.064) | 0.0375-0.346 | 0.011* |
|  |  |  | 3 vs 1 | 0.168 (0.065) | 0.012-0.323 | 0.032* |
|  |  |  | 3 vs 2 | 0.014 (0.090) | -0.208-0.236 | 0.988 |
|  |  |  | 3 vs 1 | 0.160 (0.089) | -0.059-0.378 | 0.188 |
| TTO |  |  |  |  |  |  |
| Cervicitis | 8.95 | 0.0003* | 2 vs 1 | 0.086 (0.046) | -0.024-0.196 | 0.156 |
|  |  |  | 3 vs 2 | 0.129 (0.0508) | 0.008-0.250 | 0.034* |
|  |  |  | 3 vs 1 | 0.215 (0.0508) | 0.094-0.336 | 0.000* |
| Severe PID | 5.87 | 0.0039* | 2 vs 1 | -0.062 (0.054) | -0.190 -0.066 | 0.486 |
|  |  |  | 3 vs 2 | -0.139 (0.059) | -0.280-0.002 | 0.054 |
|  |  |  | 3 vs 1 | -0.201 (0.059) | -0.342- -0.060 | 0.003* |
| Ectopic pregnancy | 8.49 | 0.0004* | 2 vs 1 | -0.123 (0.052) | -0.246 – 0.0003 | 0.051 |
|  |  |  | 3 vs 2 | 0.233 (0.058) | 0.097 -0.368 | 0.000* |
|  |  |  | 3 vs 1 | 0.110 (0.058) | -0.026 – 0.245 | 0.137 |
| Chronic pelvic pain | 7.94 | 0.0006* | 2 vs 1 | -0.072 (0.058) | -0.211-0.067 | 0.43 |
|  |  |  | 3 vs 2 | 0.252 (0.064) | 0.0998 -0.405 | 0.000* |
|  |  |  | 3 vs 1 | 0.180 (0.064) | 0.028-0.333 | 0.016* |
| TFI | 9.10 | 0.0002* | 2 vs 1 | -0.062 (0.068) | -0.224-0.100 | 0.637 |
|  |  |  | 3 vs 2 | 0.309 (0.075) | 0.131-0.487 | 0.000* |
|  |  |  | 3 vs 1 | 0.247 (0.075) | 0.069-0.425 | 0.004* |

\*P < 0.05 df= degrees of freedom; Std Err = standard Error; C.I = 95% confidence interval; NS = not significant
\*df varies for the VAS because each respondent valued 3 out of 5 THS.

### Other Analysis

The linear regression which tested whether repondents’ VAS and TTO results were dependant on age-group or gender, established that neither gender nor age could predict the values assigned to all health-states when all other variables such as ethnicity and STI history were held constant. Pearson’s correlation which tested the relationship between participants VAS scores and chained TTO results found weak positive correlations between the results for cervicitis (r:0.20), mild PID (r:0.16) and ectopic pregnancy (r:0.24) and weak negative correlations for severe PID (r:-0.12) and pre-term delivery (r:-0.02). There were no statistically significant (p < 0.05) associations as assessed using the two techniques.

## DISCUSSION

### Principal Findings

The study provides estimates of the HSUVs associated with chlamydia using direct valuation methods. The VAS and TTO (conventional and chained) techniques were applied to individuals from three diverse population groups. The modal age-group for the participants was 20-24 years. Just over half of the respondents (51%) were of White ethnicity. This is representative of the 2011 census figures [42] that show that 58% of individuals in the study area are White.

For chained TTO and VAS, ectopic pregnancy (anchor health-state) was valued as the worst THS while cervicitis was deemed the least bad THS. Chronic pelvic pain was assigned the lowest value of any of the health states (0.40 (SD-0.27)).

### Feasibility

The feasibility of conventional methods has been widely discussed [43-45], but few studies have provided such evidence for chained approaches [35,46]. The typical assumption is that the chained approach is difficult and cognitively burdensome for respondents [27,28], thus the techniques are rarely used. However, based on findings from this study (low rates of missing values and non-responses, and reasonable completion time) chained TTO should *be* deemed as feasible [35,39,40]. Caution should be applied in interpreting this result as this study was on a relatively educated population with a small sample size and may not reflect the general population.

The mean completion time in the three study populations ranged from 21-31 minutes. Completion time in our study is in keeping with previous studies, as evidenced by a published review of the literature [25] which reported an average completion time of 25 minutes.

### Time Trade-off Utilities

A broad range of HSUVs was obtained for the mean TTO utilities. The values estimated are lower than similar HSUVs typically used to populate previous decision-analytic models in cost-effectiveness studies [8]. Usually, HSUVs are used to calculate QALYs [3] and variations in these values have implications for the results of economic evaluations. Where HSUVs are low, this is an indication that the condition has a severe impact of QoL. There is a lot of scope in these cases for improvements QoL, as a result of treatment or prevention (as opposed to when the impact of the condition is small and there are minor decrements in utility). Opinions on the cost-effectiveness of chlamydia screening in England differ [47-49]. Based on our findings, and holding all other assumptions constant, if the results of this study were used to populate these models, the implication is that the results would suggest screening is more cost-effective than previously thought. However, the ultimate decision on this depends on the threshold value set for the ICER and the probability assigned in any cost-effectiveness model to the likelihood of these sequelae occurring, which is typically based on assumptions.

### VAS scores vs. TTO utilities

The VAS scores were consistently lower than TTO utilities. This result is consistent with well-documented findings that TTO elicited HSUVs are higher than values derived from VAS [45,50,51]. Unlike TTO and SG, VAS is not choice-based and does not incorporate risk and time preferences [52]. The differences in the framing of the valuation questions could explain why, typically, VAS results are lower than TTO results [53].

### Anchor health-state

In line with the decision of the experts who, before HSUV elicitation, had selected ectopic pregnancy as the worst (anchor) THS, a majority of the respondents felt this health-state was the worst THS. The selection of an appropriate anchor is a common challenge in chained TTO [25] with studies using diverse approaches [54,55]. Often in studies where the anchor state was pre-selected, some respondents preferred the anchor health-state to another THS [34,56,57]. In this study, a few respondents deemed severe PID as worse than the anchor health-state, a finding in keeping with a previous study [31]. In studies where a sizeable number of respondents had such different preferences, HSUVs for those respondents could not be calculated [35,58] as there are no guidelines. However, in such circumstances, we adopted the procedure proposed by Jansen [36].

A downward bias was observed in the anchor HSUVs. While some studies have suggested that the values assigned to health-states decrease with increasing health-states duration [59], others have shown that they are unaffected by duration [56,60]. We applied 10-year duration for the valuation of the anchor health-state. Johnston and colleagues [58] used 12-month duration for breast cancer-related THSs and similarly got lower than expected utilities for the anchor health-state. Locadia *et al*., [56] also observed that utilities obtained in Stage-II exhibited a downward bias. Both authors [56, 58] reported that respondents’ low valuation could have been influenced by the imminence of death after a short time (12 months) or the use of death as a lower anchor.

For this study, despite using a longer duration for the anchor, a downward bias was observed, supporting suggestions that respondents are sensitive to the use of dead as a lower anchor [58]. Additionally, the anchor HSUVs were on a par with utilities for the CHSs (valued with conventional TTO); further supporting suggestions that the use of dead as a lower anchor was a more likely determinant of respondents’ values than duration, as a downward bias of conventional TTO compared to chained TTO has been reported [56]. Alternately, the inclusion of the possibility of death in the health-states description for the anchor health-state could have influenced repondents’ valuations.

### Strengths and limitations

The main strength of this study is that it is the first to derive HSUVs for chlamydia using appropriate methodological approaches. Recognising that the use of standard instruments such as EQ-5D to facilitate indirect methods are not appropriate for some diseases, the study used the direct approach. The study started from first principles to develop health-state descriptions that depicted as accurately as possible the acute and chronic effects of chlamydia. This study is also the first in this disease area to use appropriate techniques targeted at specific health-states, for example, conventional TTO for CHSs and chained TTO for THSs.

Furthermore, we showed that within the same study, different approaches could be used for different health-states. Previous studies have typically used conventional methods for both chronic and temporary health states, but this is inappropriate and may have provided unreliable results. Many studies have avoided chained approaches due to their perceived complexity, but this study has shown that chained TTO is feasible.

In developing the TTO procedure [26] and the self-completed TTO version[32], provision was made for the valuation of health-states considered worse than dead. However, we didn’t consider this provision, as it could serve as an additional burden for participants. Moreover, based on previous studies [30,31], such a scenario was deemed as unlikely. The interviews were conducted by one researcher and the responses could have been biased due to interviewer’s effect.

Finally, from a purely statistical viewpoint, merging data from three different populations may be deemed inappropriate due to differences in population characteristics and methodologies [38]. However, in valuing health-states, the values of non-patients should count [1] and these populations, though diverse, represent the views of the general public. Furthermore, the merged data served to increase sample size, whilst there is transparency as to the influence of each subgroup.

### Previous research

Previous studies have typically avoided the chained approach [25]. Studies that valued health-states similar to ours, though acknowledging that some health-states were temporary [25], used conventional TTO.

### The implication for policy and practice

This study elicited HSUVs for chlamydia, an STI of public health importance. These utilities will be useful in economic evaluations requiring QALY estimates and inform policy-making on the cost-effectiveness of chlamydia interventions.

### Recommendation for future research

There is a need for more studies exploring other techniques such as chained SG, to test the reliability of the HSUVs derived. Further research assessing the impact of duration on the utility of the THS would provide greater insight into the derived HSUVs.

## CONCLUSION

This study has derived HSUVs for the health-states associated with chlamydia and its sequelae. The study has overcome methodological challenges associated with defining health-state descriptions and selection, and duration of the anchor health-state. The utilities will be useful for calculating QALYs within CUAs. While the disutilities associated with ‘chronic pelvic pain’, ‘ectopic pregnancy’ and ‘severe PID’ were substantial, the disutilities associated with the other health-states were less considerable. This could have implications for the cost-effectiveness of screening and other chlamydia interventions.

## Data Availability

Non applicable

## Notes

### Competing Interest Statement

The authors have declared no competing interest.

### Author Declarations

Ethical approval for all University-based participants (Study 1 and Study 3)was obtained from the University of Birmingham Science, Technology, Engineering and Mathematics Ethical Review Committee (ERN_16-1522) Ethical approval for recruitment of Study 2 participants was obtained from the National Health Service (NHS) Health Research Authority (IRAS ID-194331) and the Research Governance Office of the University Hospitals Birmingham NHS Foundation Trust (RRK 6073)

