## Supplementary Information 3 for "The valuation of outcomes for the temporary and chronic health states associated with Chlamydia trachomatis infection"

### Measuring the Impacts

# of

### Sexually Transmitted Infections

#### Questionnaire Booklet

| | |
|---|---|
| **Date** | **/ /2017** |
| **Participant ID** | |

#### Introduction

Welcome and thank you for choosing to take part in this study. Before you start, here is a brief introduction about what will happen as you complete this questionnaire.

This questionnaire includes some questions on how you would feel about being in some imaginary health states. If you come to any question that you do not want to answer, please let me know so I can skip it and go on to the next question.

*Thank you.*

This questionnaire includes descriptions of health states (scenarios) that females with sexually transmitted infections (STIs) may experience.

There are five sections (Sections A to E) in this questionnaire. The first four sections (A to D) involve different ways of measuring health states. The last section (E) includes some questions about you, simply to help with the analysis.

For all the sections, please read the instructions carefully as this is different for each section.

### Section A

In this section, some health state descriptions are presented. Please go through each of the health states to be valued, one at a time.

To help you say how good or bad a health state is, we have a line (like a thermometer) on page 5. On this line, the best health state you can imagine is marked 10 and the worst health state you can imagine is marked 0.

**I would like you to indicate on this scale how good or bad each of these scenarios would be for you.**

For each health state, please draw a line from the box on the opposite page to whichever point on the scale indicates how good or bad the scenario would be for you.

An example is shown on pages 4 and 5. The example does not relate to sexual health as we do not want it to influence your own responses.

Please complete the questionnaire in the same way for the health states given on pages 6 to 11.

### Example A

#### Health state description.

#### Imagine a 20-year-old female in the scenario below:

| | **Health state** |
|---|---|
| **Symptoms** | -Headache.<br>-Fever.<br>-Pain in her tummy/stomach. |
| **Impact on health-related Quality of Life** | -Feels very ill and unable to do her usual activities.<br>-Will definitely need to see a doctor.<br>-Feels worried about the nature of the illness. |
| **Treatment** | -Will be given injections for some days. |
| **Implications** | -If not treated, it could progress to cause complications.<br>-Will probably return to her usual health once fully treated.<br>-There is a small chance of the condition spreading to the lungs and causing pneumonia. |

#### Example A

If you feel that the health state was only slightly better than the worst imaginable health state, you might mark it like this:

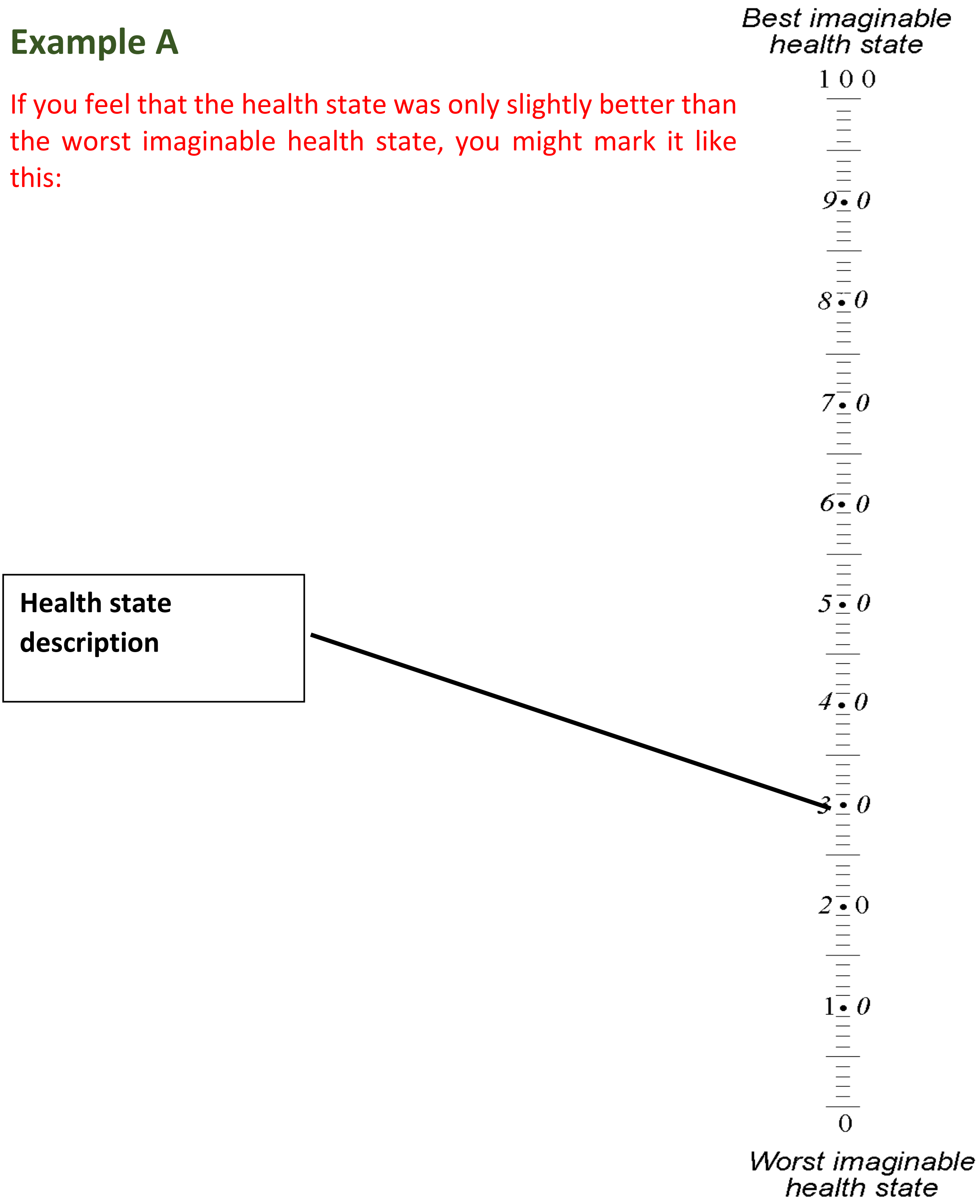


Now please draw a line in the same way for the following health states.

### Health state description.

### Imagine a 20-year-old female in the scenario below:

| | **Health state 1** |
|---|---|
| **Symptoms** | -A change in her vaginal discharge.<br>-Some bleeding or spotting after sex or in between periods.<br>-No fever. |
| **Impact on health-related Quality of life** | -May feel a bit ill but can do her usual activities.<br>-Will definitely need to see a doctor and get tested.<br>-Partner will need notifying, testing and maybe treating.<br>-Feels anxious about going to a clinic and about the nature of the disease.<br>-Feel guilty/angry about giving/catching the illness to/from her partner. |
| **Treatment** | -Will take antibiotics for a week or less. |
| **Implications** | -If not treated, could resolve itself spontaneously or progress to cause complications.<br>-She will probably return to her usual health once fully treated but could catch the infection again if she has sex with someone who is infected (an untreated partner or a new partner).<br>-The infection may spread to the uterus and cause problems with having a baby in the future. |

#### Question 1

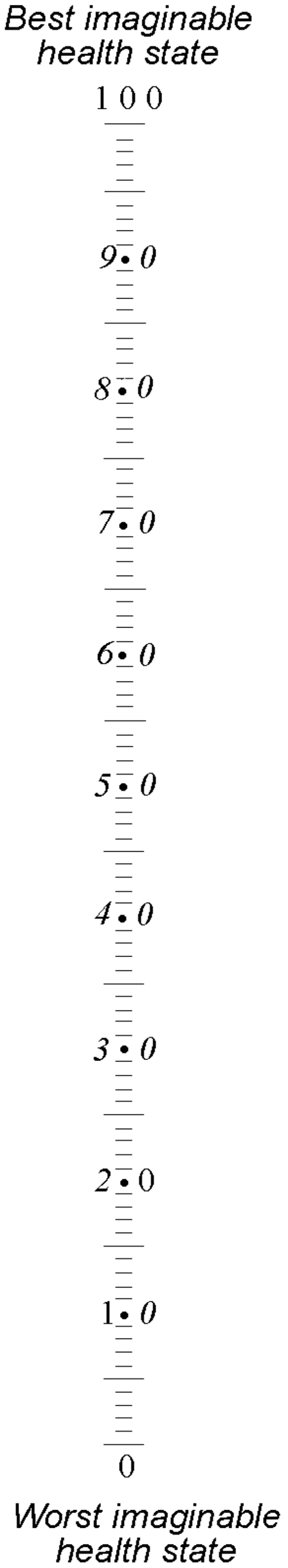


**Health state 1**

Please draw a line to show what you think about this health state compared to the best or worst imaginable health states.

### Health state description.

### Imagine a 25-year-old female in the scenario below:

| | **Health state 2** |
|---|---|
| **Symptoms** | -A change in her vaginal discharge.<br>-Pain in her lower abdomen (tummy or stomach).<br>-Some bleeding after sex.<br>-Pain during sex, severe enough to make her need to stop having sex. |
| **Impact on health-related Quality of Life** | -Will feel a bit ill and unable to do her usual activities.<br>-Will need to see a doctor and get tested.<br>-Partner will need notifying, testing and maybe treating.<br>-Feels anxious about the nature of the illness.<br>-Feels guilty/angry about giving/catching the illness to/from her partner.<br>-Has concerns about being able to have a baby in the future. |
| **Treatment** | -Will need two different types of antibiotics and these might have some side effects.<br>-May need pain killers.<br>-Will need a pregnancy test.<br>-Will need to be seen again by the doctor for follow-up. |
| **Implications** | -Could acquire the infection again if she has sex with an untreated partner.<br>-Could have a pregnancy outside the womb later.<br>-She may not be able to have a baby in the future. |

#### Question 2

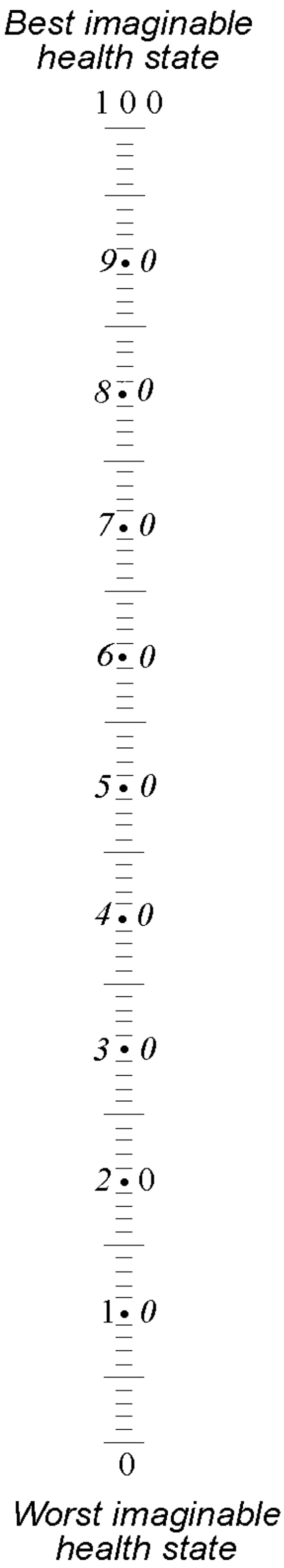


**Health state 2**

Please draw a line to show what you think about this health state compared to the best or worst imaginable health states.

### Health state description.

### Imagine a 30-year-old female in the scenario below:

| | **Health state 3** |
|---|---|
| **Symptoms** | -A change in vaginal discharge.<br>-Really severe pain in her lower abdomen (tummy or stomach).<br>-Bleeding after sex.<br>-Severe pain during sex and will definitely have to stop having sex.<br>-Fever and sometimes nausea. |
| **Impact on health-related Quality of Life** | -Will feel quite ill and hardly able to get out of bed.<br>-Will need to go to A&E or see a GP quickly.<br>-Partner will need notifying, testing and maybe treating.<br>-Feels anxious about the nature of the illness.<br>-Feels guilty/angry about giving/getting the illness to/from her partner.<br>-Has real concerns about being able to have a baby in the future. |
| **Treatment** | -Will require stay(s) in the hospital.<br>-Will need 2 or more antibiotics with bad side effects (antibiotics may be given through the vein at first).<br>-May need pain killers.<br>-Will need a pregnancy test.<br>-May require an invasive test (in which objects will be put inside the body) and an operation. |
| **Implications** | -Could acquire the infection again if she has sex with an untreated partner.<br>-She may have a pregnancy outside the womb later.<br>-There is a high possibility that she may not be able to have a baby in the future. |

#### Question 3

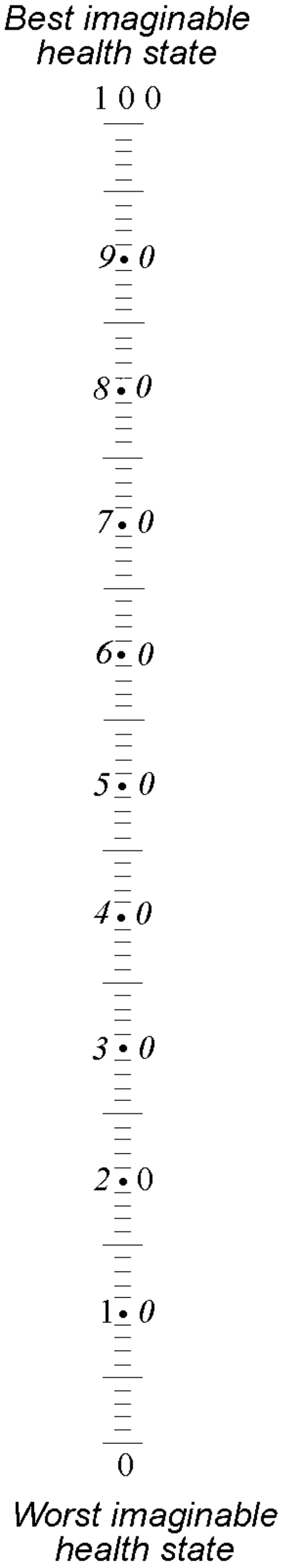


**Health state 3**

Please draw a line to show what you think about this health state compared to the best or worst imaginable health states.

**Please do not write on this page**

#### Section B

In this section, you will be asked to make choices between different health states.

The health state descriptions are labelled as Scenario A and Scenario B. Note that these health state descriptions are different in every question.

For Scenario A, we would like you to imagine that you will live for 30 years in the health state described and then you will die.

For Scenario B, we would like you to imagine that you will live for **less** than 30 years in the health state described and then you will die.

We would like you to consider whether you would prefer to live a shorter life but in better health (as in Scenario B), or a longer life in worse health by indicating how many years in Scenario B would be equal to or the same as thirty years in Scenario A.

An example is shown on pages 14 and 15. The example does not relate to sexual health as we do not want it to influence your own responses.

For each question on pages 16 to 19 please write your answer as shown in the example.

### Example B

| Scenario A | Health state: 20-year-old female |
|---|---|
| **Symptoms** | -Headache.<br>-Fever.<br>-Pain in her tummy/stomach. |
| **Impact on health-related Quality of Life** | -Feels very ill and unable to do her usual activities.<br>-Will definitely need to see a doctor.<br>-Feels worried about the nature of the illness. |
| **Treatment** | -Will be given injections for some days. |
| **Implications** | -If not treated, it could progress to cause complications.<br>-Will probably return to her usual health once fully treated.<br>-A small chance of the condition spreading to the lungs and causing pneumonia. |

| Scenario B | Full health state: A 20-year-old female |
|---|---|
| **Symptoms** | -Normal vaginal discharge.<br>-No pain in her tummy or during sex.<br>-No bleeding or spotting after sex or in between periods. |
| **Impact on health-related Quality of Life** | -Feels able do her usual activities.<br>-Does not need to see a doctor.<br>-Able to have healthy sex life.<br>-Feels happy and fulfilled about her sexual health. |
| **Treatment** | -Will not need to take any medications. |
| **Implications** | -No problem with having a baby in the future. |

#### Example B

Please put an "A" against all cases where you are CONFIDENT that you would choose Scenario A.

Please put a "B" against all cases where you are CONFIDENT that you would choose Scenario B.

Please put a "=" against all cases where you cannot choose between Scenario A and Scenario B.

In this example, the person completing the questionnaire feels that 8 years in Scenario B is the same as 30 years in Scenario A.

| Scenario A | | Scenario B |
|---|---|---|
| 30 years | B | 30 years |
| 30 years | B | 29 years |
| 30 years | B | 28 years |
| 30 years | B | 27 years |
| 30 years | B | 26 years |
| 30 years | B | 25 years |
| 30 years | B | 20 years |
| 30 years | B | 15 years |
| 30 years | B | 10 years |
| 30 years | = | 8 years |
| 30 years | A | 6 years |
| 30 years | A | 4 years |
| 30 years | A | 2 years |
| 30 years | A | 1 year |
| 30 years | A | 0 years |

Now do the exercise for the following health states.

### Question 1

| Scenario A | Health state 6: 35-year-old female |
|---|---|
| **Symptoms** | -Continuous pain in her lower abdomen.<br>-Severe pain during sex and this pain will definitely make her need to stop having sex. |
| **Impact on health-related Quality of Life** | -Pain interferes with her family and social relationships.<br>-Unable to go to the gym or do other activities.<br>-Often feels downhearted and unhappy.<br>-Feels anxious about her condition.<br>-Longs to feel fulfilled and have more energy. |
| **Treatment** | -Will need a pregnancy test.<br>-May need an invasive test (in which objects will be put inside the body).<br>-May require an operation.<br>-Will need to take pain relievers and antibiotics. |
| **Implications** | -The pain may slowly go away as time goes on but it could also stay the same. |

| Scenario B | Full health state: A 35-year-old female |
|---|---|
| **Symptoms** | -Normal vaginal discharge.<br>-No pain in her tummy or during sex.<br>-No bleeding or spotting after sex or in between periods. |
| **Impact on health-related Quality of Life** | -Feels able do her usual activities.<br>-Does not need to see a doctor.<br>-Able to have healthy sex life.<br>-Feels happy and fulfilled about her sexual health. |
| **Treatment** | -Will not need to take any medications. |
| **Implications** | -No problem with having a baby in the future. |

#### Question 1

Please put an “A” against all cases where you are CONFIDENT that you would choose Scenario A.

Please put a “B” against all cases where you are CONFIDENT that you would choose Scenario B.

Please put a “=” against all cases where you cannot choose between Scenario A and Scenario B

| Scenario A | | Scenario B |
|---|---|---|
| 30 years | | 30 years |
| 30 years | | 29 years |
| 30 years | | 28 years |
| 30 years | | 27 years |
| 30 years | | 26 years |
| 30 years | | 25 years |
| 30 years | | 20 years |
| 30 years | | 15 years |
| 30 years | | 10 years |
| 30 years | | 8 years |
| 30 years | | 6 years |
| 30 years | | 4 years |
| 30 years | | 2 years |
| 30 years | | 1 year |
| 30 years | | 0 years |

#### Question 2

| Scenario A | Health state 7: A 35-year-old female |
|---|---|
| **Symptoms** | -Trying to get pregnant (unsuccessfully) for over one year.<br>-May have chronic pain in her pelvis. |
| **Impact on health-related Quality of Life** | -Feels stressed because she really wants to have children as soon as possible.<br>-Feels worthless and hopeless.<br>-Feels unhappy and less satisfied in her relationship.<br>-Will need to see a doctor for assessment. |
| **Treatment** | -May need to undergo some tests some of which are quite invasive.<br>-May need to have an operation.<br>-May have to undergo repeated courses of expensive treatment (not covered by the NHS), which may or may not correct the condition. |
| **Implications** | -There is a chance that the situation may not be resolved. |

| Scenario B | Full health state: A 35-year-old female |
|---|---|
| **Symptoms** | -Normal vaginal discharge.<br>-No pain in her tummy or during sex.<br>-No bleeding or spotting after sex or in between periods. |
| **Impact on health-related Quality of Life** | -Feels able do her usual activities.<br>-Does not need to see a doctor.<br>-Able to have healthy sex life.<br>-Feels happy and fulfilled about her sexual health. |
| **Treatment** | -Will not need to take any medications. |
| **Implications** | -No problem with having a baby in the future. |

#### Question 2

Please put an “A” against all cases where you are CONFIDENT that you would choose Scenario A.

Please put a “B” against all cases where you are CONFIDENT that you would choose Scenario B.

Please put a “=” against all cases where you cannot choose between Scenario A and Scenario B

| Scenario A | | Scenario B |
|---|---|---|
| 30 years | | 30 years |
| 30 years | | 29 years |
| 30 years | | 28 years |
| 30 years | | 27 years |
| 30 years | | 26 years |
| 30 years | | 25 years |
| 30 years | | 20 years |
| 30 years | | 15 years |
| 30 years | | 10 years |
| 30 years | | 8 years |
| 30 years | | 6 years |
| 30 years | | 4 years |
| 30 years | | 2 years |
| 30 years | | 1 year |
| 30 years | | 0 years |

**Please do not write on this page**

### Section C

Now you will be asked to make choices between some of the scenarios that you saw in the first section.

Scenario A and Scenario B indicate health state descriptions. Note that these health state descriptions differ in every question.

In Scenario A, we would like you to imagine that you will spend 12 months (one year) in the health state described and then you will return to full health.

For Scenario B, we would like you to imagine that you spend **less** than 12 months (one year) in the health state described and then you will return to full health.

We would like you to consider how many months in Scenario B would be equivalent to one year in Scenario A.

An example is shown on pages 22 and 23. The example does not relate to sexual health as we do not want it to influence your own responses.

For each question on pages 24 to 31 please write your answer as shown in the example.

#### Example C

| Scenario A | Health state: 20-year-old female |
|---|---|
| **Symptoms** | -Headache.<br>-Fever.<br>-Pain in her tummy/stomach. |
| **Impact on health-related Quality of Life** | -Feels very ill and unable to do her usual activities.<br>-Will definitely need to see a doctor.<br>-Feels worried about the nature of the illness. |
| **Treatment** | -Will be given injections for some days. |
| **Implications** | -If not treated, it could progress to cause complications.<br>-Will probably return to her usual health once fully treated.<br>-A small chance of the condition spreading to the lungs and causing pneumonia. |

| Scenario B | Health state: A 20-year-old female |
|---|---|
| **Symptoms** | -Normal vaginal discharge.<br>-No pain in her tummy or during sex.<br>-No bleeding or spotting after sex or in between periods. |
| **Impact on health-related Quality of Life** | -Feels able do her usual activities.<br>-Does not need to see a doctor.<br>-Able to have healthy sex life.<br>-Feels happy and fulfilled about her sexual health. |
| **Treatment** | -Will not need to take any medications. |
| **Implications** | -No problem with having a baby in the future. |

#### Example C

Please put an "A" against all cases where you are CONFIDENT that you would choose Scenario A.

Please put a "B" against all cases where you are CONFIDENT that you would choose Scenario B.

Please put a "=" against all cases where you cannot choose between Scenario A and Scenario B.

In this example, the person completing the questionnaire feels that 9 months in Scenario B is the same as 12 months in Scenario A.

| Scenario A | | Scenario B |
|---|---|---|
| 12 months | A | 12 months |
| 12 months | A | 11 months |
| 12 months | A | 10 months |
| 12 months | = | 9 months |
| 12 months | B | 8 months |
| 12 months | B | 7 months |
| 12 months | B | 6 months |
| 12 months | B | 5 months |
| 12 months | B | 4 months |
| 12 months | B | 3 months |
| 12 months | B | 2 months |
| 12 months | B | 1 months |
| 12 months | B | 0 months |

Now do the exercise for the following health states.

### Question 1

| Scenario A | Health state 1: A 20-year-old female |
|---|---|
| **Symptoms** | -A change in her vaginal discharge.<br>-Some bleeding or spotting after sex or in between periods.<br>-No fever. |
| **Impact on health-related Quality of life** | -May feel a bit ill but can do her usual activities.<br>-Will definitely need to see a doctor and get tested.<br>-Partner will need notifying, testing and maybe treating.<br>-Feels anxious about going to a clinic and about the nature of the disease.<br>-Feel guilty/angry about giving /catching the illness to/from her partner. |
| **Treatment** | -Will take antibiotics for a week or less. |
| **Implications** | -If not treated, could resolve itself spontaneously or progress to cause complications.<br>-She will probably return to her usual health once fully treated but could catch the infection again if she has sex with someone who is infected (an untreated partner or a new partner).<br>-There is a small chance of the infection spreading to the uterus and causing problems with having a baby in the future. |

| Scenario B | Health state 4: A 20-year old female |
|---|---|
| **Symptoms** | -Lower abdominal cramp or pain, often localised to one part of the stomach.<br>-May miss a period.<br>-Light vaginal bleeding.<br>-May have diarrhoea.<br>-May feel dizzy. |
| **Impact on health-relate Quality of Life** | -Will need to go quickly to the emergency services.<br>-May need to be treated to remove the cause of the illness.<br>-May need to go to the hospital several times.<br>-May need several blood tests.<br>-Feels anxious about the diagnosis.<br>-Feels worried that she could die and about being able to have a baby in the future. |
| **Treatment** | -May need to have a minor operation.<br>-May need to get 1 or more injections in the arm.<br>-A small chance of the uterine tube bursting and will require an operation at once to remove the uterine tube. |
| **Implications** | - There is a possibility that the uterine tube will need to be removed<br>-She may not be able to have a baby in the future.<br>-There is a very small chance of death.<br>-There is a possibility of the recurrence of the illness. |

#### Question 1

Please put an “A” against all cases where you are CONFIDENT that you would choose Scenario A.

Please put a “B” against all cases where you are CONFIDENT that you would choose Scenario B.

Please put a “=” against all cases where you cannot choose between Scenario A and Scenario B

| Scenario A | | Scenario B |
|---|---|---|
| 12 months | | 12 months |
| 12 months | | 11 months |
| 12 months | | 10 months |
| 12 months | | 9 months |
| 12 months | | 8 months |
| 12 months | | 7 months |
| 12 months | | 6 months |
| 12 months | | 5 months |
| 12 months | | 4 months |
| 12 months | | 3 months |
| 12 months | | 2 months |
| 12 months | | 1 month |
| 12 months | | 0 months |

### Question 2

| Scenario A | Health state 2: 25-year-old female |
|---|---|
| **Symptoms** | -A change in her vaginal discharge.<br>-Pain in her lower abdomen (tummy or stomach).<br>-Some bleeding after sex.<br>-Pain during sex, severe enough to make her need to stop having sex. |
| **Impact on health-related Quality of Life** | -Will feel a bit ill and unable to do her usual activities.<br>-Will need to see a doctor and get tested.<br>-Partner will need notifying, testing and maybe treating.<br>-Feels anxious about the nature of the illness.<br>-Feels guilty/angry about giving/catching the illness to/from her partner.<br>-Has concerns about being able to have a baby in the future |
| **Treatment** | -Will need two different types of antibiotics and these might have some side effects.<br>-May need pain killers.<br>-Will need a pregnancy test.<br>-Will need to be seen again by the doctor for follow-up. |
| **Implications** | -Could acquire the infection again if she has sex with an untreated partner.<br>-She could have a pregnancy outside the womb later.<br>-She may not be able to have a baby in the future. |

| Scenario B | Health state 4: A 20-year-old female |
|---|---|
| **Symptoms** | -Lower abdominal cramp or pain, often localised to a part of the stomach.<br>-May miss a period.<br>-Light vaginal bleeding.<br>-May have diarrhoea.<br>-May feel dizzy. |
| **Impact on health-related Quality of Life** | -Will need to go quickly to the emergency services.<br>-May need to be treated to remove the cause of the illness.<br>-May need to go to the hospital several times.<br>-May need several blood tests.<br>-Feels anxious about the diagnosis.<br>-Feels worried that she could die and about being able to have a baby in the future. |
| **Treatment** | -May need to have a minor operation.<br>-May need to get 1 or more injections in the arm.<br>-A small chance of the uterine tube bursting and will require an operation at once to remove the uterine tube. |
| **Implications** | - There is a possibility that the uterine tube will need to be removed<br>-She may not be able to have a baby in the future.<br>-There is a very small chance of death.<br>-There is a possibility of the recurrence of the illness. |

#### Question 2

Please put an “A” against all cases where you are CONFIDENT that you would choose Scenario A.

Please put a “B” against all cases where you are CONFIDENT that you would choose Scenario B.

Please put a “=” against all cases where you cannot choose between Scenario A and Scenario B

| Scenario A | | Scenario B |
|---|---|---|
| 12 months | | 12 months |
| 12 months | | 11 months |
| 12 months | | 10 months |
| 12 months | | 9 months |
| 12 months | | 8 months |
| 12 months | | 7 months |
| 12 months | | 6 months |
| 12 months | | 5 months |
| 12 months | | 4 months |
| 12 months | | 3 months |
| 12 months | | 2 months |
| 12 months | | 1 month |
| 12 months | | 0 months |

#### Question 3

| Scenario A | Health state 3: 30-year-old female |
|---|---|
| Symptoms | -A change in vaginal discharge.<br>-Really severe pain in her lower abdomen (tummy or stomach).<br>-Bleeding after sex.<br>-Severe pain during sex and will definitely have to stop having sex.<br>-Fever and sometimes nausea. |
| Impact on health-related Quality of Life | -Will feel quite ill and can hardly get out of bed.<br>-Will need to go to A&E or see a GP quickly.<br>-Partner will need notifying, testing and maybe treating.<br>-Feels anxious about the nature of the illness.<br>-Feels guilty/angry about giving/getting the illness to/from her partner.<br>-Has real concerns about being able to have a baby in the future. |
| Treatment | -Will require stay(s) in the hospital.<br>-Will need 2 or more antibiotics with bad side effects (antibiotics may be given through the veins at first).<br>-May need pain killers.<br>-Will need a pregnancy test.<br>-May require an invasive test and an operation. |
| Implications | -Could acquire the infection again if she has sex with an untreated partner.<br>-She could have a pregnancy outside the womb later.<br>-There is a high possibility that she may not be able to have a baby in the future. |

| Scenario B | Health state 4: A 20-year old female |
|---|---|
| Symptoms | -Lower abdominal cramp or pain, often localised to a part of the stomach.<br>-May miss a period.<br>-Light vaginal bleeding.<br>-May have diarrhoea.<br>-May feel dizzy. |
| Impact on health-relate Quality of Life | -Will need to go quickly to the emergency services.<br>-May need to be treated to remove the cause of the illness.<br>-May need to go to the hospital several times.<br>-May need several blood tests.<br>-Feels anxious about the diagnosis.<br>-Feels worried that she could die and about being able to have a baby in the future. |
| Treatment | -May need to have a minor operation.<br>-May need to get 1 or more injections in the arm.<br>-A small chance of the uterine tube bursting and will require an operation at once to remove the uterine tube. |
| Implications | -There is a possibility that the uterine tube will need to be removed<br>-She may not be able to have a baby in the future.<br>-There is a very small chance of death.<br>-There is a possibility of the recurrence of the illness. |

#### Question 3

Please put an "A" against all cases where you are CONFIDENT that you would choose Scenario A.

Please put a "B" against all cases where you are CONFIDENT that you would choose Scenario B.

Please put a "=" against all cases where you cannot choose between Scenario A and Scenario B

| **Scenario A** | | **Scenario B** |
|---|---|---|
| 12 months | | 12 months |
| 12 months | | 11 months |
| 12 months | | 10 months |
| 12 months | | 9 months |
| 12 months | | 8 months |
| 12 months | | 7 months |
| 12 months | | 6 months |
| 12 months | | 5 months |
| 12 months | | 4 months |
| 12 months | | 3 months |
| 12 months | | 2 months |
| 12 months | | 1 month |
| 12 months | | 0 months |

#### Question 4

| Scenario A | Health state 5: 20-year-old pregnant female |
|---|---|
| **Symptoms** | -Is 12 to 14 weeks away from her due-date.<br>-Pain in the lower back and cramps in the lower tummy.<br>-Pain and spasm come and go but do not stop.<br>-She might wet her pants. |
| **Impact on health-relate Quality of Life** | -Will need to go to the emergency services at once.<br>-Feels anxious about her condition.<br>-Feels worried about the outcome for the baby. |
| **Treatment** | -Will need to stay in the hospital for several weeks.<br>-Will receive fluids and other medicines through the vein.<br>-Will be given injections. |
| **Implications** | -After treatment, will return to normal health.<br>-There will be need for frequent hospital appointments in the future for the baby. |

| Scenario B | Health state 4: A 20-year old female |
|---|---|
| **Symptoms** | -Lower abdominal cramp or pain, often localised to a part of the stomach.<br>-May miss a period.<br>-Light vaginal bleeding.<br>-May have diarrhoea.<br>-May feel dizzy. |
| **Impact on health-relate Quality of Life** | -Will need to go quickly to the emergency services.<br>-May need to be treated to remove the cause of the illness.<br>-May need to go to the hospital several times.<br>-May need several blood tests.<br>-Feels anxious about the diagnosis.<br>-Feels worried that she could die and about being able to have a baby in the future. |
| **Treatment** | -May need to have a minor operation.<br>-May need to get 1 or more injections in the arm.<br>-A small chance of the uterine tube bursting and will require an operation at once to remove the uterine tube. |
| **Implications** | - There is a possibility that the uterine tube will need to be removed<br>-She may not be able to have a baby in the future.<br>-There is a very small chance of death.<br>-There is a possibility of the recurrence of the illness. |

#### Question 4

Please put an “A” against all cases where you are CONFIDENT that you would choose Scenario A.

Please put a “B” against all cases where you are CONFIDENT that you would choose Scenario B.

Please put a “=” against all cases where you cannot choose between Scenario A and Scenario B

| Scenario A | | Scenario B |
|---|---|---|
| 12 months | | 12 months |
| 12 months | | 11 months |
| 12 months | | 10 months |
| 12 months | | 9 months |
| 12 months | | 8 months |
| 12 months | | 7 months |
| 12 months | | 6 months |
| 12 months | | 5 months |
| 12 months | | 4 months |
| 12 months | | 3 months |
| 12 months | | 2 months |
| 12 months | | 1 month |
| 12 months | | 0 months |

**Please do not write on this page**

#### Section D

In this section, Scenario A and Scenario B indicate different health state descriptions. Note that these health state descriptions differ in every question.

For Scenario A, we would like you to imagine that you will live for 10 years in the health state described and then you will die.

For Scenario B, we would like you to imagine that you will live for **less** than 10 years in the health state described and then you will die.

We would like you to consider whether you might prefer to live a shorter life but in better health or a longer life in worse health by indicating how many years in Scenario B would be the same to you as to 10 years in Scenario A.

An example is shown on pages 34 and 35. The example does not relate to sexual health as we do not want it to influence your own responses.

For the question on pages 36 to 37 please write your answer as shown in the example.

#### Example D

| Scenario A | Health state: A 20-year-old female |
|---|---|
| Symptoms | -Headache.<br>-Fever.<br>-Pain in her tummy/stomach. |
| Impact on health-related Quality of Life | -Feels very ill and unable to do her usual activities.<br>-Will definitely need to see a doctor.<br>-Feels worried about the nature of the illness. |
| Treatment | -Will be given injections for some days. |
| Implications | - If not treated, it could progress to cause complications.<br>-Will probably return to her usual health once fully treated.<br>-A small chance of the condition spreading to the lungs and causing pneumonia. |

| Scenario B | Full health state: A 20-year-old female |
|---|---|
| Symptoms | -Normal vaginal discharge.<br>-No pain in her tummy or during sex.<br>-No bleeding or spotting after sex or in between periods. |
| Impact on health-related Quality of Life | -Feels able do her usual activities.<br>-Does not need to see a doctor.<br>-Able to have healthy sex life.<br>-Feels happy and fulfilled about her sexual health. |
| Treatment | -Will not need to take any medications. |
| Implications | -No problem with having a baby in the future. |

#### Example D

Please put an “A” against all cases where you are CONFIDENT that you would choose Scenario A.

Please put a “B” against all cases where you are CONFIDENT that you would choose Scenario B.

Please put a “=” against all cases where you cannot choose between Scenario A and Scenario B

In this example, the person completing the questionnaire feels that 7 years in Scenario B is the same as 10 years in Scenario A.

| Scenario A | | Scenario B |
|---|---|---|
| 10 years | B | 10 years |
| 10 years | B | 9 years |
| 10 years | B | 8 years |
| 10 years | = | 7 years |
| 10 years | A | 6 years |
| 10 years | A | 5 years |
| 10 years | A | 4 years |
| 10 years | A | 3 years |
| 10 years | A | 2 years |
| 10 years | A | 1 year |
| 10 years | A | 0 years |

Now do the exercise for the following health states.

### Question 1

| Scenario A | Health state 4<br>20-year-old female |
|---|---|
| **Symptoms** | -Lower abdominal cramp or pain, often localised to one part of the stomach.<br>-May miss a period.<br>-Light vaginal bleeding.<br>-May have diarrhoea<br>-May feel dizzy. |
| **Impact on health-relate Quality of Life** | -Will need to go quickly to the emergency services.<br>-May need to be treated to remove the cause of the illness.<br>-May need to go to the hospital several times.<br>-May need several blood tests.<br>-Feels anxious about the diagnosis.<br>-Feels worried that she could die and about being able to have a baby in the future. |
| **Treatment** | -May need to have a minor operation.<br>-May need to get 1 or more injections in the arm.<br>-A small chance of the uterine tube bursting and will require an operation at once to remove the uterine tube. |
| **Implications** | -There is a possibility that the uterine tube will need to be removed<br>-She may not be able to have a baby in the future.<br>-There is a very small chance of death.<br>-There is a possibility of the recurrence of the illness. |

| Scenario B | Full health state: A 20-year-old female |
|---|---|
| **Symptoms** | -Normal vaginal discharge.<br>-No pain in her tummy or during sex.<br>-No bleeding or spotting after sex or in between periods. |
| **Impact on health-related Quality of Life** | -Feels able do her usual activities.<br>-Does not need to see a doctor.<br>-Able to have healthy sex life.<br>-Feels happy and fulfilled about her sexual health. |
| **Treatment** | -Will not need to take any medications. |
| **Implications** | -No problems with having a baby in the future. |

#### Question 1

Please put an “A” against all cases where you are CONFIDENT that you would choose Scenario A.

Please put a “B” against all cases where you are CONFIDENT that you would choose Scenario B.

Please put a “=” against all cases where you cannot choose between Scenario A and Scenario B

| Scenario A | | Scenario B |
|---|---|---|
| 10 years | | 10 years |
| 10 years | | 9 years |
| 10 years | | 8 years |
| 10 years | | 7 years |
| 10 years | | 6 years |
| 10 years | | 5 years |
| 10 years | | 4 years |
| 10 years | | 3 years |
| 10 years | | 2 years |
| 10 years | | 1 year |
| 10 years | | 0 years |

### Section E
### Questionnaire on background information

*Here are some general questions about you. Before you start, I will like to remind you that if you come to any question that you do not want to answer, please skip it and go onto the next question. Thank you.*

*First are some questions about your background.*

1. I identify my gender as

☐ Male.
☐ Female.
☐ Trans*.
☐ Other (*please specify*) ……………………………..

2. What is your ethnicity? *(Please tick one box only).*

☐ White British.
☐ White, other background.
☐ Asian / Asian British.
☐ Black British.
☐ Black, other background.
☐ Mixed ethnic group.
☐ Other ethnic group.
☐ Prefer not to say.

3. What was your age at your last birthday? (*Please tick one box only*).

☐ 16-17.
☐ 18-19.
☐ 20-24.
☐ 25-34.
☐ Over 34.

4. Which of the following best describes your main activity? *(Please tick one box only).*

☐ In employment or self-employment.
☐ Student.
☐ Seeking work.
☐ Other *(please specify)* ………………………………………….....

*The next questions are about infections that can be transmitted by sex. Please answer, even if you have never had an infection that was transmitted by sex.*

5. How would you describe your relationship status?

☐ Not in a relationship.
☐ In a steady relationship.
☐ In a casual relationship.
☐ Prefer not to say.

6. How many sex partners have you had in the last year?

☐ None.
☐ 1.
☐ 2-5.
☐ 6-10.
☐ > 10.
☐ Prefer not to say.

7. Have you ever been told by your doctor or any health care professional that you had any of the following?

☐ Chlamydia.
☐ Gonorrhoea.
☐ Genital warts.
☐ Pelvic Inflammatory disease (PID) (*women only*).
☐ Vaginal thrush (Candida) (*women only*).
☐ Epididymitis (*men only*).
☐ Others (*please specify*) ……………………………………….
☐ Yes, but cannot remember which.
☐ No.
☐ Prefer not to say.

8. In the last year have you been tested for Chlamydia?

☐ Yes.
☐ No**.**
☐ Prefer not to say.

9. Why were you last tested for Chlamydia?

- ☐ I had symptoms.
- ☐ My partner had symptoms.
- ☐ I was notified because a partner was diagnosed with Chlamydia.
- ☐ I wanted a general sexual health check-up.
- ☐ Check up after previous positive test.
- ☐ I had no symptoms but I was worried about the risk of Chlamydia.
- ☐ I was offered a routine test.
- ☐ I have never been tested.
- ☐ Other (*please specify*) ……………………………..

Thank you for your time.
