## Supplementary Information 1_2 for "The valuation of outcomes for the temporary and chronic health states associated with Chlamydia trachomatis infection"

### Supplemental Documents

#### S1: Health-State description development

First, depictions of health-states associated with chlamydia and its complications were drafted based on literature. Next, experts in Sexual Health, Obstetrics/Gynaecology and Health Economics (with experience in economic evaluation and outcome measures) were requested to review the initial descriptions. These Experts (6) were mostly form the researcher's institution while five were from other institutions.

| **Sexual health panel** | | |
|---|---|---|
| Initial | Designation | Institution |
| CE | Professor of Sexual Health and HIV | External |
| JS | Clinical Champion/ NCSP | External |
| JR | Professor of Sexual Health and HIV | External |
| MS | Research Fellow in Infection and Immunity | External |
| NL | Professor of Epidemiology and Public Health | External |
| **Obstetrics and Gynaecology panel** | | |
| Initial | Designation | |
| JG | Professor of Obstetrics and Gynaecology | Internal |
| AC | Professor of Gynaecology | Internal |
| JC | Professor of Obstetrics and Gynaecology | Internal |
| **Health Economics team** | | |
| Initial | Designation | |
| TR | Professor of Health Economics | Internal |
| LJ | Lecturer in Health Economics | Internal |
| PK | Research Fellow in Health Economics | Internal |
| **Internal –** From Researcher's Institution | | |
| **Questionnaire Review** | | |
| This was a pragmatic and informal review with colleagues and their children if their children were over 18 years old to see if questions were clear. The results were destroyed and not relevant as the assessment was on clarity of questions. The respondents had nothing to do with the main study. | | |

This led to a second draft of the health-state descriptions that was used to develop a questionnaire booklet.The questionnaire booklet presented the health states in the context of the TTO task. The overall task (using the questionnaire booklet) was piloted with a convenience sample of eight young adults aged 16-24 years. This was to assess the questionnaire for understanding and feasibility. Based on the responses from the pilot, some grammatical corrections were made to the health descriptions, along with modifications to the format of the TTO table.

### S2: Example of a health-state description

| | **Health-state 1** |
|---|---|
| **Symptoms** | -A change in her vaginal discharge.<br>-Some bleeding or spotting after sex or in between periods.<br>-No fever. |
| **Impact on health-related quality of life** | -May feel a bit ill but can do her usual activities.<br>-Will definitely need to see a doctor and get tested.<br>-Partner will need notifying, testing and maybe treating.<br>-Feels anxious about going to a clinic and about the nature of the disease.<br>-Feel guilty/angry about giving/catching the illness to/from her partner. |
| **Treatment** | -Will take antibiotics for a week or less. |
| **Implications** | -If not treated, could resolve itself spontaneously or progress to cause complications.<br>-She will probably return to her usual health once fully treated but could catch the infection again if she has sex with someone who is infected (an untreated partner or a new partner).<br>-The infection may spread to the uterus and cause problems with having a baby in the future. |
